# The external validity of machine learning-based prediction scores from hematological parameters of COVID-19: A study using hospital records from Brazil, Italy, and Western Europe

**DOI:** 10.1101/2023.03.07.23286949

**Authors:** Ali Akbar Safdari, Chanda Sai Keshav, Deepanshu Mody, Kshitij Verma, Utsav Kaushal, Vaadeendra Kumar Burra, Sibnath Ray, Debashree Bandyopadhyay

## Abstract

**Background:** The COVID-19 pandemic is the deadliest threat to humankind caused by the SARS-COV-2 virus in recent times. The gold standard for its detection, quantitative Real-Time Polymerase Chain Reaction (qRT-PCR), has several limitations regarding experimental handling, expense, and time. While the hematochemical values of routine blood tests have been reported as a faster and cheaper alternative, the external validity of the model on a diverse population has yet to be thoroughly investigated. Here we studied the external validity of machine learning-based prediction scores from hematological parameters recorded in Brazil, Italy, and Western Europe.

**Methods and Findings:** The publicly available hematological records (raw sample size (n) = 195554) from hospitals of three different territories, Brazil, Italy, and Western Europe, were preprocessed to develop the training, testing, and prediction cohorts for ML models. A total of eight (sub)datasets were trained on seven different ML classifiers. The XGBoost classifier performed consistently better on all the datasets producing eight different models. The working models include a set of either four or fourteen hematological parameters. The internal performances of the XGBoost models (AUC scores range from 84% to 97%) were superior to the ML models reported in the literature for a few datasets (AUC scores range from 84% to 87%). The external performance (AUC score) was 86% when the model was trained and tested on fourteen hematological parameters obtained from the same country (Brazil) but on independent datasets. However, the external performances were reduced when tested across the populations; 69% when trained on datasets from Italy (n=1736) and tested on datasets from Brazil (n=602)) and 65%, when trained on datasets from Italy and tested on datasets from Western Europe (n=1587)) respectively.

**Conclusion:** For the first time, this report showed that the models trained and tested on the same population but on separate records produced reasonably accurate results. The study promises the confidence of these models trained and tested within the same populations and has the potential application to extend those to other demographic locations. Both four- and fourteen-parameter models are publicly available; https://covipred.bits-hyderabad.ac.in/home

**Author Summary:** COVID-19 has posed the deadliest threat to the human population in the 21^st^ century. Timely detection of the disease could save more lives. The RT-PCR test is considered the gold standard for COVID-19 detection. However, there are several limitations of the technique that suggests developing an alternate detection protocol that would be efficient, fast, and cheap. Among several other alternate detection techniques, hematology based Machine-Learning (ML) prediction is one. All the hematology-based predictions reported so far in the literature were only internally validated. Considering the need to develop an alternate protocol for rapid, near-accurate, and cheaper COVID-19 detection techniques, we aim to externally validate the hematology-based ML prediction. Here external validation indicates use of two independent datasets for model training and testing, in contrast to internal validation where the same dataset splits into train and test sets. We have integrated published clinical records from Brazil, Italy, and West Europe hospitals. Internal ML model performances are superior compared to those reported in literature. The external model performances were equivalent to the internal performances when trained and tested on the same population. However, the external performances were inferior when train and test sets were from different populations. The results promise the utility of these models on the same populations. However, it also warns to train the model on one population and test it on another. The outcome of this work has the potential for an initial screen of COVID-19 based on hematological parameters before qRT-PCR tests.

## Introduction

The COVID-19 infection has posed the deadliest threat to the health of the human population in the 21^st^ century. Likely, the danger is far from over concerning the emerging variants of COVID-19, such as alpha (B.1.1.7), beta (B.1.351), gamma (P.1), delta (B.1.617.2), lambda (C.37), and omicron (B.1.1.529) ^1^, along with other frequently mutating respiratory diseases, like, influenza virus A (H1N1) ^2^. Due to the nature of the disease, timely detection of COVID-19 is of utmost importance. Hence, detection techniques play a pivotal role in its diagnosis. The gold standard of COVID-19 detection is quantitative Real-Time Polymerase-Chain-Reaction (qRT-PCR). This method has several limitations, like manual errors during sample (nasal and oral swab) collection, operational errors, etc. ^3^. Moreover, the time required for the experiment and availability of the detection kits at a mass level becomes difficult in a vast population with a large number of infections. The test is also costly for low-income groups. An accurate, rapid, and low-cost prediction strategy would supplement the initial screening, particularly in a country like India, with the second-largest population in the world.

The most common clinical feature of severe COVID-19 is pneumonia with fever, cough, fatigue, headache, diarrhea, hypoxia, and dyspnoea. The latest variant, omicron, has some common symptoms with the earlier SARS-COV-2 strains, although with lesser severity due to mild infection in the lower respiratory tract and reduced probability of hospitalization^1^. In the case of mild COVID-19 infection, either no (asymptotic) or only mild pneumonia is observed. In moderate infection, dyspnoea, hypoxia, and lung injury may occur. In severe infection, respiratory failure to multi-organ failure occurs. In brief, severe cases of COVID-19 can lead to a systemic infection affecting almost all of the major organ systems. As a result, patients of COVID-19 exhibit a wide range of hematologic abnormalities that changes with disease progression, severity, and mortality ^4^. For example, the white blood cells sense and respond to the microbial threats ^5^; blood platelet expression and platelet counts are altered ^6 7^ - platelet hyperactivity was demonstrated as one of the unique features of COVID-19 infection ^8^. Hence, a complete blood count (CBC) could serve as a biomarker for COVID-19. Screening the COVID-19 infection in terms of CBC has been attempted by various research groups worldwide, ^9 10 11 12 13 14^.

Some of these research groups used machine learning (ML) approaches to exploit the CBC parameters from a specific population for disease prediction; the Area Under Curve (AUC) performance ranges from 84% to 87% in those models. So far, no report is available to test the applicability of the hematology-based ML models across different ethnicity and populations. The combination of CBC parameters varies with ethnicity. However, some blood parameters alteration, such as lymphocytopenia^15 16^ leucopenia, and thrombocytopenia ^17 18 19^, are common due to COVID-19. In this work, we combined different hematological parameters from various populations to develop the optimal ML models and tested them on independent datasets obtained from other populations. Standardization across different ML algorithms yields eXtreme Gradient Boost (XGBoost) as the best-performing model across the datasets compared to published literature. For the first time, we report the external validity of the prediction scores trained, tested, and predicted across the populations. The models performed the best when trained and tested on the same population but on different records (datasets).

## Method

### Description of clinical datasets for training, validation, and prediction

#### Dataset 1

Dataset-1 was generated based on anonymized patient data publicly available from Hospital Israelita Albert Einstein, in São Paulo, Brazil https://www.kaggle.com/einsteindata4u/covid19. The data were recorded from February 26^th^, 2020, to March 23^rd^, 2020. The cases and controls for this dataset include the patients whose samples were collected to perform the SARS-CoV-2 qRT-PCR and additional laboratory tests during a visit to the hospital.

The initial data set consisted of 558 positive and 5086 negative cases of COVID-19. This dataset was processed to minimize the null-value columns and eliminate the negative instances with many null values. The value (xi) in each cell was pre-normalized (at the source) to a mean value (μ) of zero and a unit standard deviation (σ); this was termed as ‘normalized count’; xi’ = (xi-μ)/σ. The same normalization scheme has been used throughout the subsequent datasets. The columns with null values appearing more than 90% were dropped. The records (rows) showing positive results were retained by default, and the negative records were maintained only with more than 10% non-null entries. This processed dataset, termed dataset 1, contains thirty-seven features and 2004 records, 558 positives and 1446 negatives (Table 1). The negative to positive sample size ratio, 2.59, is four times less than that in the published model (11.51)^9^. Here ‘features’ refer to x-parameters used to train the model; the definition excludes the y-parameter, SARS-COV2 results (positive or negative). This definition is consistently used in the subsequent datasets. These thirty-seven features were categorized into four classes, namely, i) age, ii) severity of the infection, iii) hematological features, and iv) co-morbidities (Table S1).

**Table 1:** Statistics of the datasets

| Dataset | No. of entries (P+N) | No. positive cases (P) | No. of negative cases (N) | Default scale_posweight (=N/P) | No. of features used |
| --- | --- | --- | --- | --- | --- |
| 1 (i) | 2004 | 558 | 1446 | 2.59 | 37 |
| 1a | 602 | 83 | 519 | 6.25 | 18 |
| 1b | 602 | 83 | 519 | 6.25 | 14 |
| 1c | 602 | 83 | 519 | 6.25 | 4 |
| 2a (ii) | 1388 | 765 | 623 | 0.81 | 31 |
| 2b | 1736 | 816 | 920 | 1.13 | 4 |
| 3a (iii) | 5872 | 1772 | 4100 | 2.31 | 21 |
| 3b | 12105 | 8926 | 3176 | 0.356 | 14 |

I. https://www.kaggle.com/einsteindata4u/covid19
II. https://zenodo.org/record/4081318#.X4RWqdD7TIU
III. https://repositoriodatasharingfapesp.uspdigital.usp.br/

#### Dataset 1a

A subset of dataset 1 was curated with eighteen features – patient age quantile, three hospitalization conditions, namely, patients admitted to regular ward, semi-ICU, and ICU, and fourteen hematological parameters. Co-morbidities were excluded from dataset 1a. The total number of records was 602, with 83 positives and 519 negatives.

#### Dataset 1b

A subset of dataset 1 was curated based on hematological features only. Other parameters, namely, co-morbidities, patient age quantile and patient admission status, were dropped in this dataset. All features with fewer than 90% of non-null values were dropped. All the records that have 100% null values were dropped. The preprocessing resulted in a dataset of fourteen hematological features and 602 records, 83 positives, and 519 negatives. Thus, the negative-to-positive sample size ratio was 6.25.

#### Dataset 1c

A third subset of dataset 1 (dataset 1c) was curated from dataset 1b based on four blood count features (Figure 1) that have shown a higher correlation with the qRT-PCR results. The number of records, positives, and negatives are identical to dataset 1b. These four blood count features were also reported as significant for SARS-COV-2 infection in published literature^9^.

**Figure 1:**
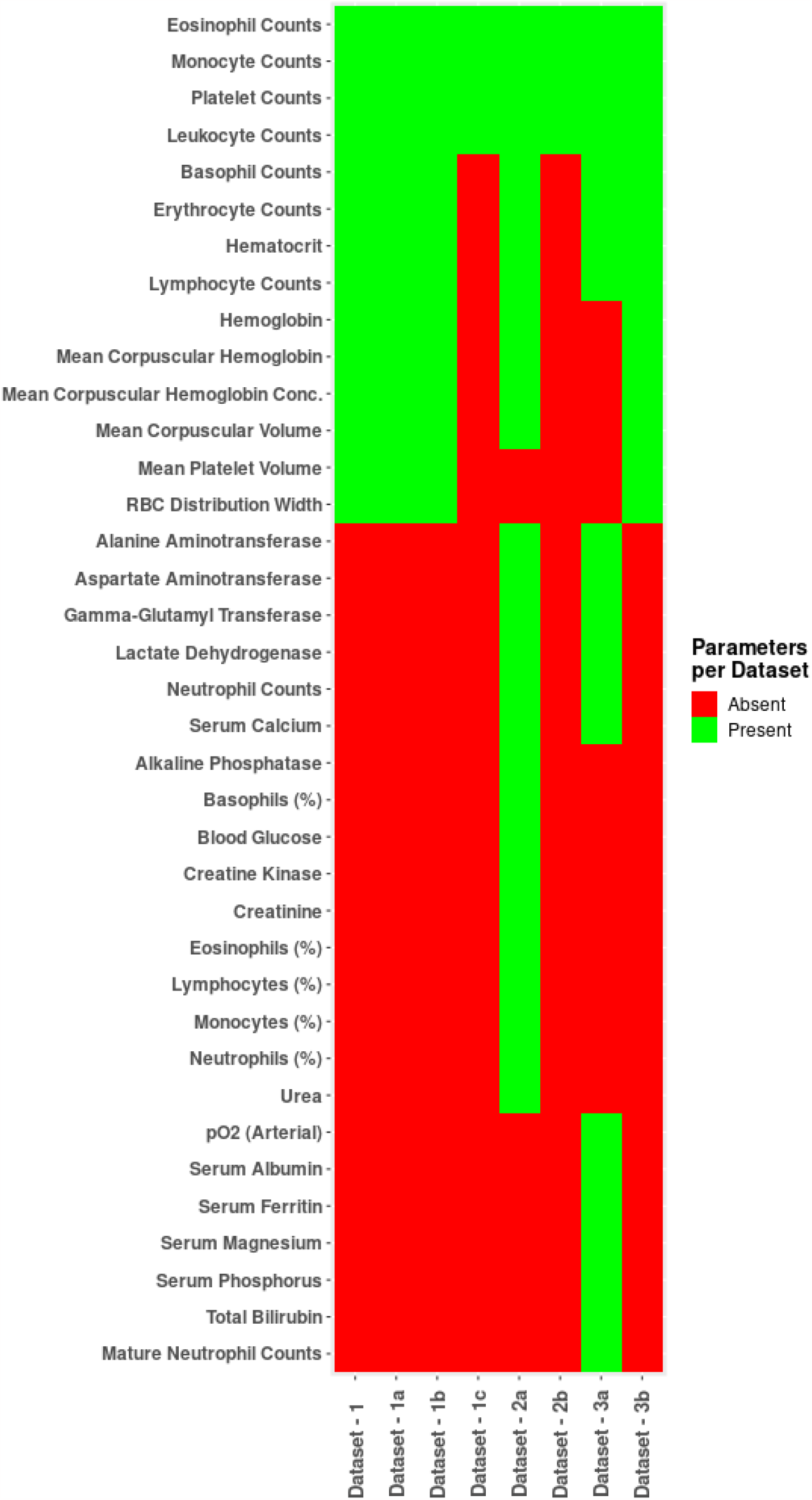
Haematological features used in different datasets. The green colour indicated the presence of a particular feature in a dataset and the red colour indicated its absence.

#### Dataset 2

This dataset was obtained from San Raphael Hospital (OSR), Italy^11^. In the original OSR dataset, there were 1736 entries with a total of 72 features, and those included 36 hematological features. The samples were collected from patients admitted to OSR from February to May 2020. Fifty-two percent of the patients were COVID-19 positive.

#### Dataset 2a

These 1736 entries were processed such that all rows (records) with more than 66% null values were dropped. The processed dataset contained 1388 records, 765 positives, and 623 negatives. This dataset includes 31 features: age, sex, a feature for suspicion (representing subjective analysis of the patient by a physician), and 28 hematological parameters (Figure 1). The ratio of negative to positive records was 0.81.

#### Dataset 2b

A subset of dataset 2 was curated with only four blood count parameters (similar to dataset 1c). No columns or rows were dropped here, as there were no rows with less than 66% null values. Dataset 2b has 1736 records, 816 positives, and 920 negatives.

#### Dataset 3

Dataset 3 was obtained from the Covid Data Sharing initiative created by a consortium led by FAPESP (Sao Paulo Research Foundation) and USP (University of Sao Paulo, Brazil). The data originated from three prominent private hospitals in Sao Paulo, Brazil - Fleury Institute, Sírio-Libanês Hospital, and Albert Einstein Hospital, from November 1^st^, 2019, to July 1^st^, 2020 (https://repositoriodatasharingfapesp.uspdigital.usp.br/). The data was anonymized from patients tested for COVID-19 (serology or RT-PCR).

The raw data obtained from the data sharing initiative had multiple rows (records) corresponding to individual patients containing different clinical features (“long-form” of the dataset). The “long form” of the dataset was converted, using an in-house python code, to the “wide form,” where one row corresponds to all the clinical features of a patient. The “wide form” of the dataset has 189227 records and 454 features. These 454 features were common, as there were duplicates in the column headers (due to different reference ranges) for some features. After deduplication, the feature number was reduced to 104.

#### Dataset 3a

The non-duplicated features were further filtered by excluding the following conditions, i) no qRT-PCR results available, ii) all the rows with more than 66% null values, and iii) the Pearson correlation of that particular feature (for the SARS-COV-2 results) less than 0.05. A total of twenty-one hematological indices (features) were identified based on the above cutoff (Figure 1). The final dataset contains 5872 records, 1772 positives, and 4100 negatives.

#### Dataset 3b

The deduplicated ‘wide form’ of the data (189227 records and 104 features) were filtered with the following conditions– a) qRT-PCR results present, b) records with null values less than 66%, and c) fourteen hematological parameters present, as in dataset 1b. The total number of records present in the dataset was 12105 records.

All the processed datasets have a 90:10 split between the training and the test data.

### Description of the clinical dataset for blind prediction

#### Western European dataset

This dataset was obtained from several hospitals in Western Europe (Table S2). The dataset includes the patients from the first day of hospitalization to nearly five weeks^13^. This published data was in the form of twenty separate tables that we merged into a single file comprising 2587 entries and thirty-seven features. According to the source authors13, there are two stages of the disease, a) early stage, from day zero through three (total of four days), and b) advanced stage, comprising all the subsequent days. This blind prediction dataset includes only four hematological parameters consistent with dataset-2b.

### Machine Learning (ML) approaches

The machine learning (ML) algorithms were implemented in Python (3.7.13) using the following libraries, Numpy (1.21.6), Pandas (1.3.5), XGBoost (0.90), Scikit-learn (1.0.2), Seaborn (0.11.2), Matplotlib (3.2.2) and Pickle 4.0 libraries.

#### Different algorithms

The algorithm primarily employed was the Extreme Gradient Boost (XGBoost) classifier that implements gradient-boosted decision trees (with enhanced speed and performance) and trains a class-weighted (or cost-sensitive) version of imbalanced classification^20^. XGBoost, a ternary classifier, considers null entries one of the classes that handle the null-entry values. Other classifiers tested on these datasets were logistic regression, Fischer linear discriminant Naïve Bayes, SVM, random forest, and K-Nearest Neighbor (KNN). Logistic regression predicts the output of a categorical dependent variable by fitting an “S” shaped logistic function that indicates two maximum values, 0 or 1. Fischer linear discriminant classifier maximizes the separation between the projected class means and minimizes the class overlap leading to well-separated classes. Naive Bayes is a classification technique based on the Bayes theorem with an assumption of independent predictors; a particular feature is independent of another feature in a class. The SVM algorithm aims to create the best line or decision boundary to segregate n-dimensional space into classes to accommodate a new data point. The best decision boundary, a hyperplane, is made based on the extreme points (vectors). Random forest is a concept of ensemble learning – a combination of multiple classifiers to solve a complex problem and improve the model performance. As the name suggests, Random Forest contains several decision trees on various subsets of the given dataset and takes the average to improve the predictive accuracy of that dataset. KNN algorithm stores all the available data and classifies a new data point based on the similarity by placing a new data point in the nearest category. Thus, new data belongs to an appropriate class.

#### Hyper-parameter used in XGBoost classifier

To normalize the imbalance in the number of negative and positive data points in the XGBoost classifier, hyper-parameter – “*scale_pos_weight*” https://xgboost.readthedocs.io/en/stable/parameter.html#parameters-for-tree-booster, was introduced. The *scale_pos_weight* value was used to scale the gradient for the positive class. For example, the “*scale_pos_weight*” = 2 indicates twice the weight of the positive class compared to the negative class. It also overcorrects the misclassification of the positive class. The loss curve (optimized to get a better model) will be affected differently in case of positive and negative entry misclassification. However, large scale_pos_weight can help the model achieve better performance for the positive class prediction (overfitting the positive class) at the cost of worse performance on the negative or both classes. Hence we have consistently considered the default *scale-pos-weight* (the ratio of numbers of negative to positive entries) throughout this report.

#### Imputation for other ML models

Unlike XGBoost, most ML algorithms cannot handle null values, thus requiring data imputation. We imputed missing values through the IterativeImputer module in the ScKit-learn package (https://scikit-learn.org/stable/modules/impute.html#multivariate-feature-imputation), which imputes values for null data points for each feature iteratively. It does so by fitting a regressor to the other feature columns (X-parameter) for records with known values of the target feature (y-parameter) and then predicts missing values of the target feature.

#### Performance metrics

The performance metrics used were accuracy, specificity, and sensitivity, defined by true positives (TP), true negatives (TN), false positives (FP), and false negatives (FN) (eq 1-3).

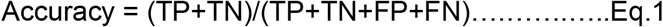

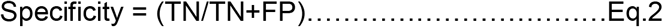

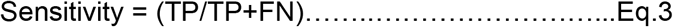

The fourth metric was the Area Under the ROC Curve (AUC). The AUC was computed from prediction scores using the roc_auc_score (https://scikit-learn.org/stable/modules/generated/sklearn.metrics.roc_auc_score.html) module of the sklearn—metrics library. A ROC curve (Receiver Operating Characteristic curve) plots the performance (True Positive Rate (TPR) versus False Positive Rate (FPR)) of a classification model at all classification thresholds. TPR is synonymous with sensitivity, also known as recall. FPR is FP/(FP + TN). AUC measures the Area under ROC (as defined by TPR versus FPR) curve from (0,0) to (1,1) along the x-axis (FPR axis). AUC ranges from 0 to 1; 0 implies a 100% wrong model, and 1 indicates a 100% correct model.

### Design of the web server

The web server hosted two different models, a four-hematological parameter model and a fourteen-hematological parameter model. The web server was developed on an HTML framework, with five working HTML files: a landing page and two pages each for each method, one for data input and the other for prediction display. The basic skeleton of the HTML files was formatted with CSS code, and these files were deployed via the python module, Flask. Python libraries, like numpy and pandas, were used to collect and process the input, with the responses generated by the XGBoost models.

## Results and Discussion

### Clinical datasets exploited for feature selection

Three independent clinical datasets (dataset 1, dataset 2, and dataset 3) were curated and processed from hospitals in Brazil and Italy (Figure 2). Hematological features were selected from these datasets based on the Pearson correlation coefficients computed between the features and the SARS-COV-2 results (positive or negative) (Figure 3).

**Figure 2:**
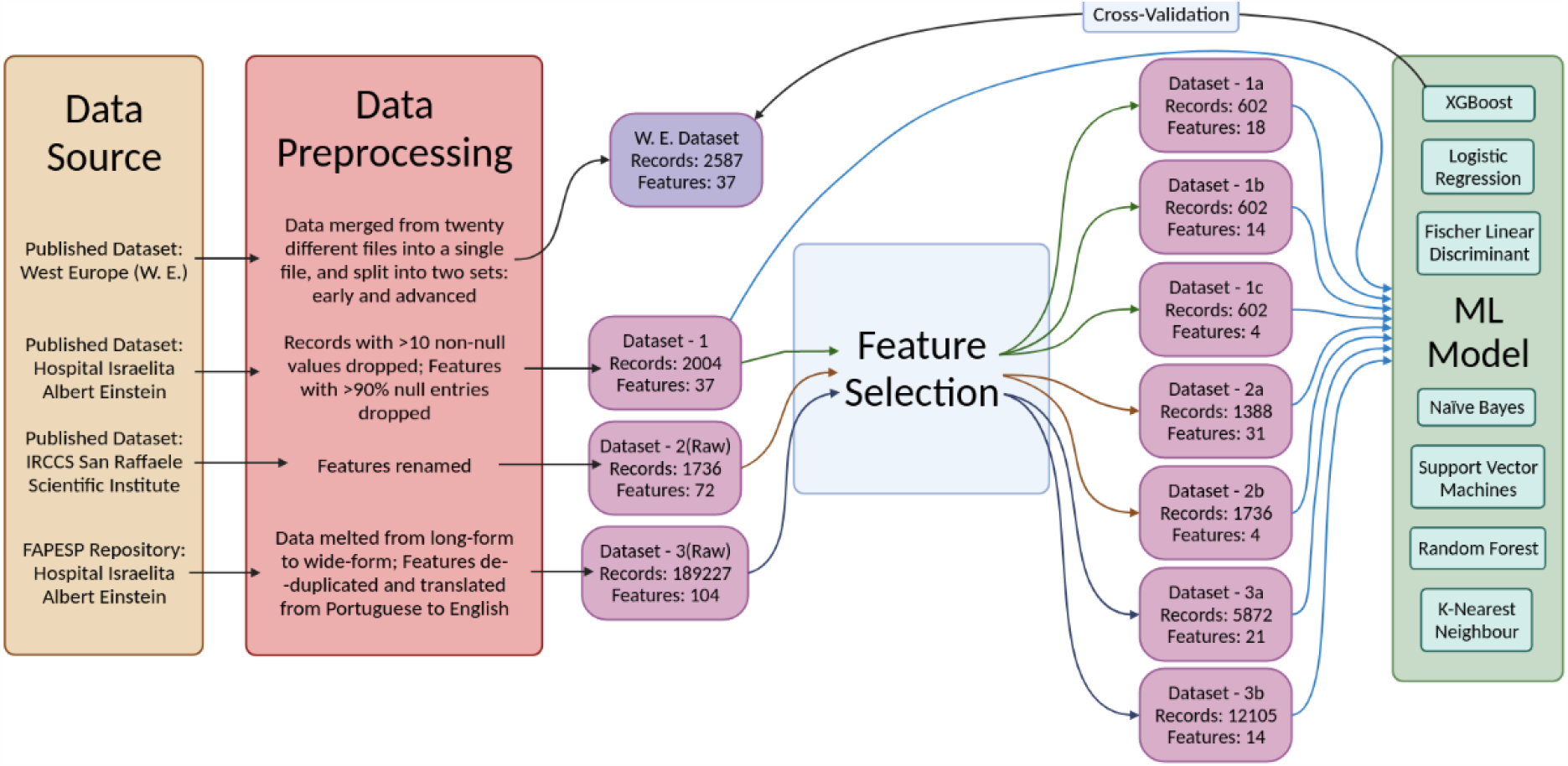
Description of data sources used for training and prediction of different ML-models based on haematological features for COVID-19 characterization

**Figure 3:**
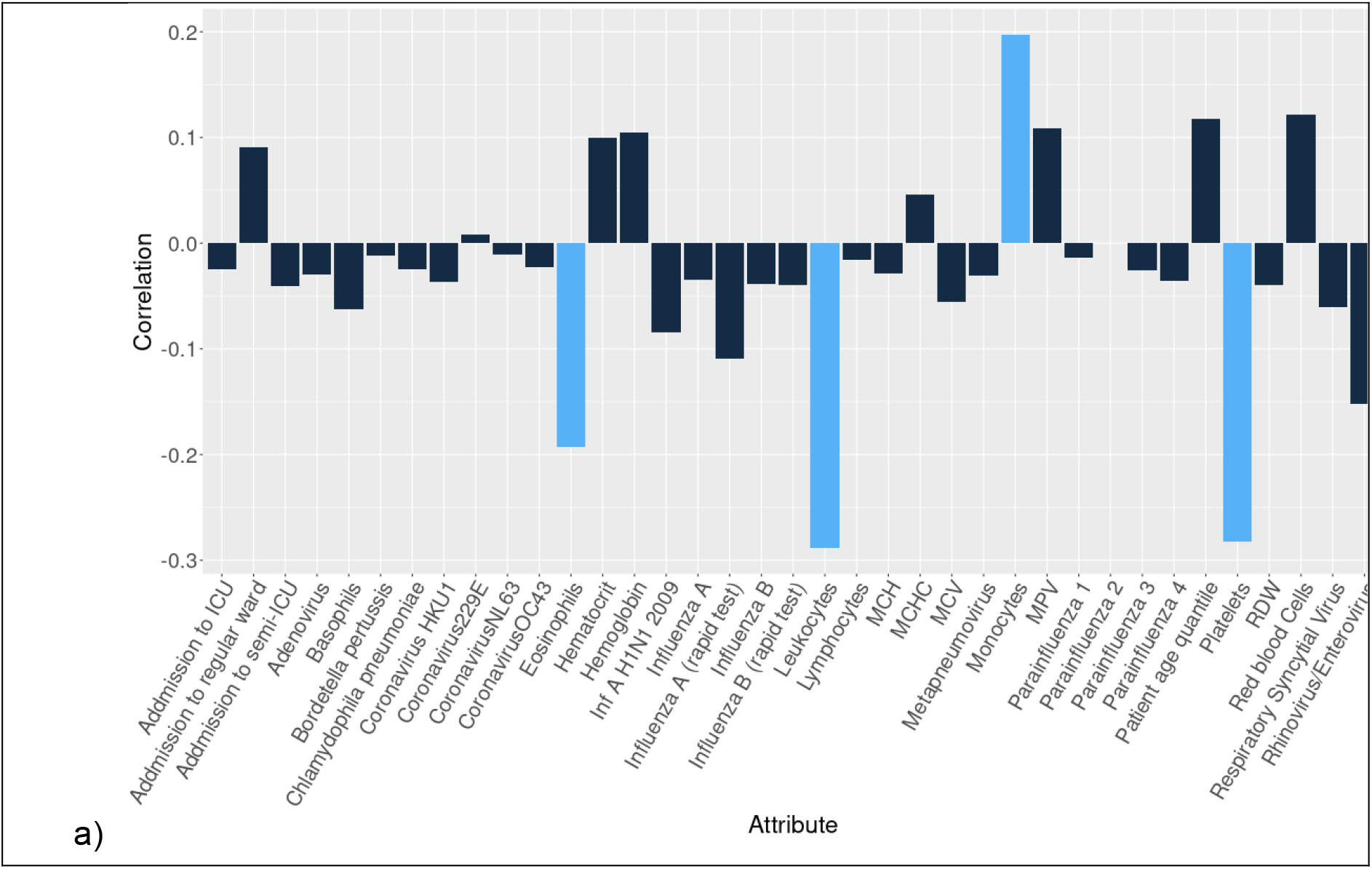
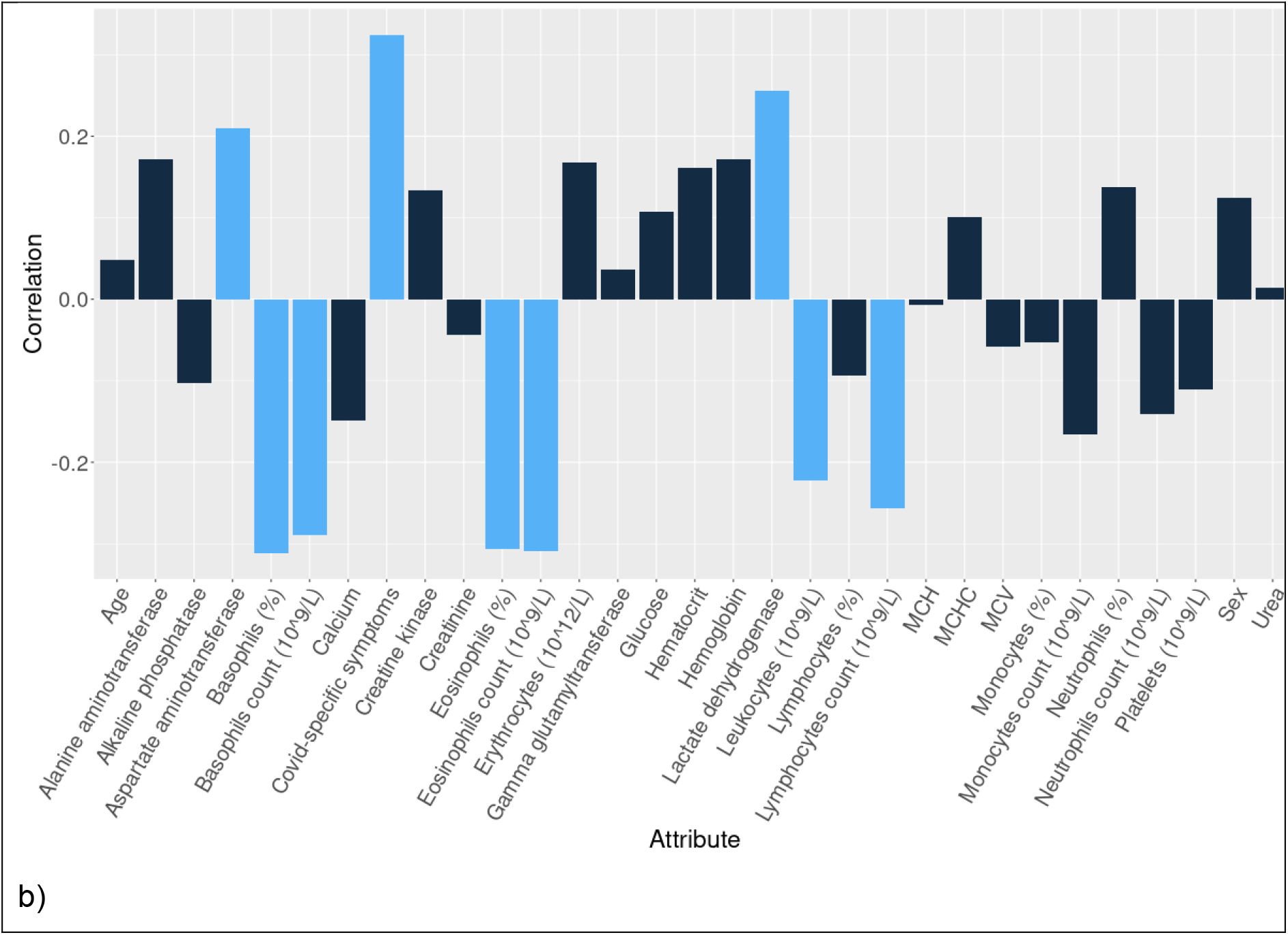
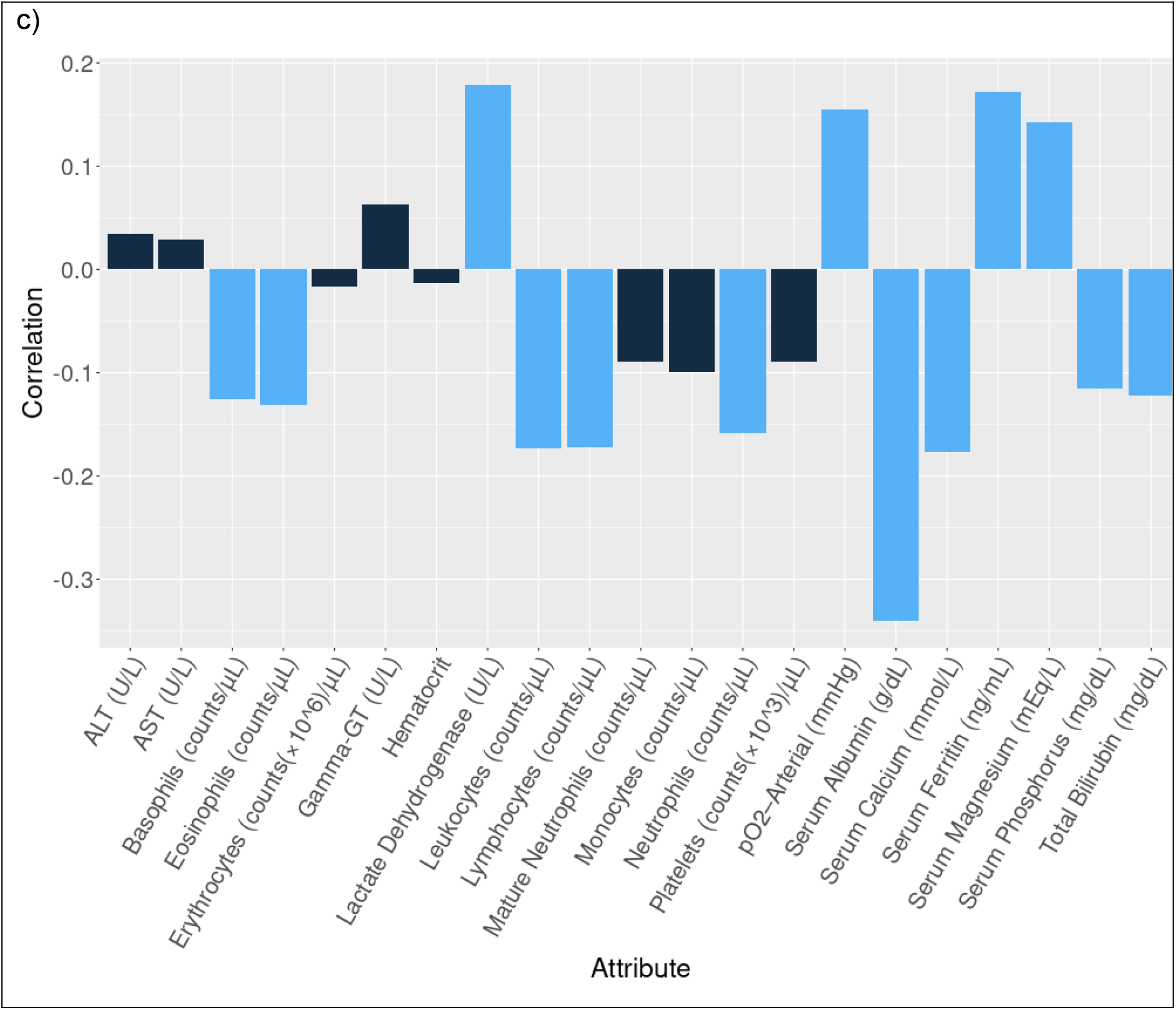
Pearson correlation coefficients between SARS-COV-2 results and individual features for a) dataset 1 b) dataset 2a and c) dataset 3a. Parameters with higher correlation (>∼±0.2) are shown in blue, remaining values in black, with an exception for dataset 3a (correlation cut off ±0.1).

For dataset-1, four features (out of thirty-seven) showed higher correlation values (cutoff value ∼±0.2) with SARS-COV-2 results. These four features were platelet counts, monocytes, eosinophils, and leukocytes (all reported in 10^9/L). Only monocytes have shown a significant increase in their values in SARS-COV-2 patients (positive correlation). The remaining parameters decreased during infection (negative correlation). Careful observation revealed that in the case of non-admitted patients, monocyte increase is maximum, suggesting that innate immunity is handling the infection. On the other hand, platelet volume (MPV) increased, and platelet counts decreased in the case of regular ward patients, clearly indicating the increase in platelet size. Thus the immune system will be affected, and the number of immune cells will decrease, justifying the negative correlation of eosinophil, leukocytes, and platelet count with SARS-COV-2 disease. The low platelet counts accounted for severe COVID-19 patients, even down in non-survivors compared to the survivors ^21^. The correlation coefficient values between SARS-COV-2 results and different features reported elsewhere were similar to this observations^11^. Hence, dataset-1c was developed on these four features.

For dataset 2a, eight features (out of twenty-eight) have shown correlation values outside the cutoff. Those features were i) aspartate aminotransferase, ii) lactate dehydrogenase, iii) leukocyte (10^9/L), iv) eosinophil (%), v) basophil (%), vi) eosinophil count, vii) lymphocyte count and viii) basophil count (all the counts in 10^9/L). Two features: Aspartate aminotransferase and lactate dehydrogenase have increased in COVID-19 patients. The remaining other hematological features decreased. In datasets 1c and 2a, there are two features, leukocyte count and eosinophil count, commonly drop with SARS-COV-2 results that presumably indicate that despite variable immune response in different populations, some hematological features are common in SARS-COV-2 disease across the populations.

For dataset 3a, four parameters, lactate dehydrogenase, partial oxygen pressure in the artery, serum ferritin, and serum magnesium, have a higher positive correlation (>0.1) with SARS-COV-2 results. Whereas basophil, eosinophil, leukocyte, and lymphocyte counts have a higher negative correlation (<-0.1) with SARS-COV-2 results. In datasets 1c, 2a, and 3a, two clinical features, leukocyte count and eosinophil count, were common.

### Comparative performances of seven different ML models on current datasets

Eight datasets (Figure 2) from three primary datasets, 1, 2, and 3, were derived based on either higher correlation with SARS-COV-2 results or to make parity (in terms of the number of features) with other datasets. The overall statistics of these eight datasets are shown (Table 1). Different ML models were trained on these datasets. The performances were measured using the receiver operating characteristic (ROC) curves (Figure 4). XGBoost outperformed other methods for all datasets except dataset 1a. The internal evaluation showed that the XGBoost model outperformed all the datasets when all four performance metrics, namely, accuracy, sensitivity, specificity, and AUC scores, were considered together (Table S3).

**Figure 4:**
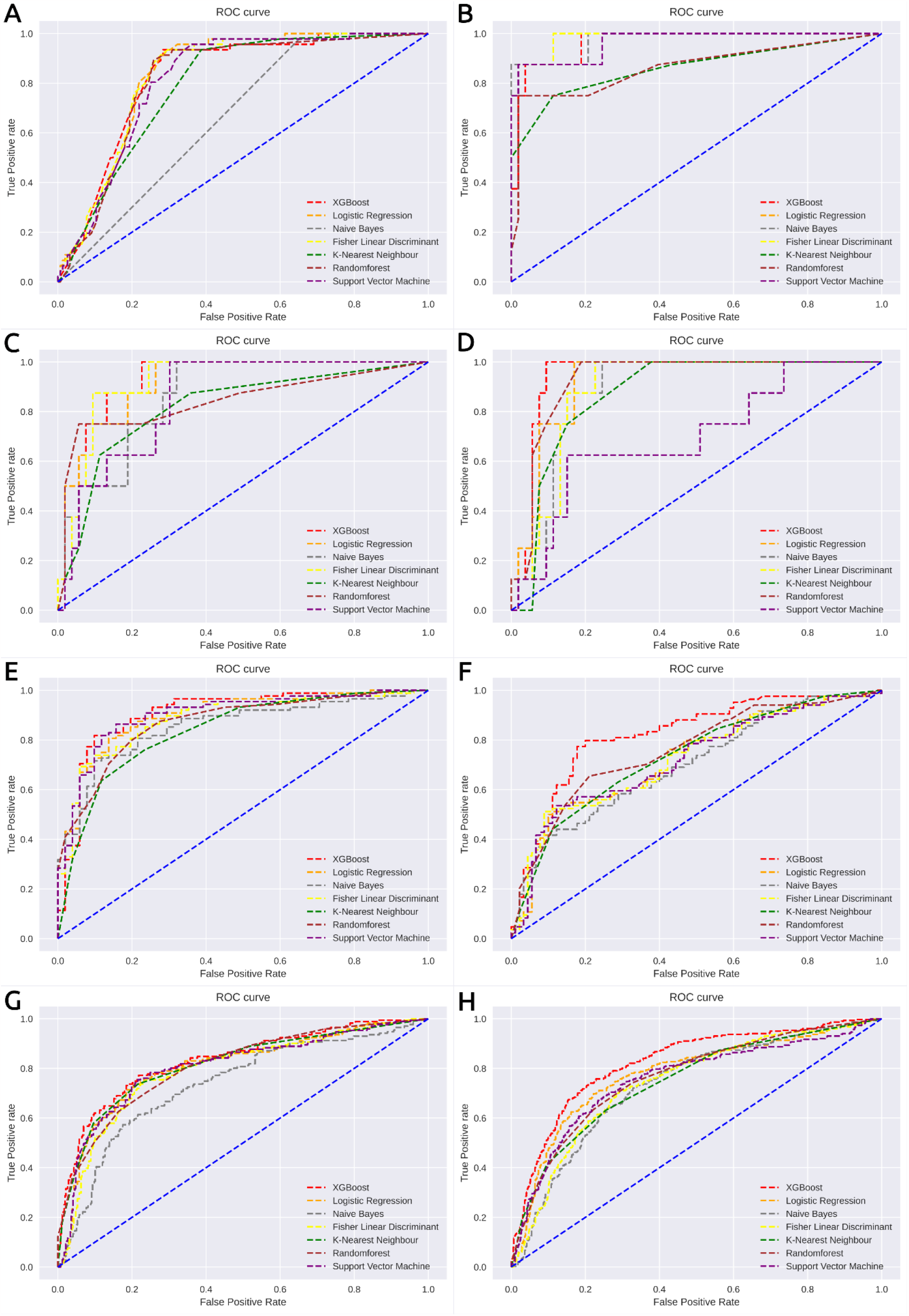
Receiver Operating Characteristics Curves (ROC) across different ML models for a) Dataset 1 b) Dataset 1a, c) Dataset 1b, d) Dataset 1c, e) Dataset 2a, f) Dataset 2b, g) Dataset 3a and h) Dataset 3b

Datasets-1 and 1c have shown optimal performances (AUC scores 0.94 and 0.97, respectively) in all four metrics. For dataset 1c, sensitivity was observed as 1.0, indicating 100% correct prediction of True Positive (TP) values, presumably, due to overcorrection of the TP values in a small dataset (n=602) with a low population of positives (n=83), leading to large *scale-pos-weight* of 6.25. As mentioned in the method section, large *scale-pos-weight* improves the performance of the positive class prediction at the cost of the negative class prediction. The XGBoost models, when compared with the other models, in terms of all the metrics, the notable observation was low sensitivity values for dataset 1 (small sample size, n=602) and allied subsets (datasets, 1a to 1c) for almost all the models except Naïve Bayes classifier. The low sensitivity values for datasets 1b and 1c are presumably attributed to the smaller size and shallow positive populations in those datasets. Most likely, the XGBoost, being a ternary classifier, can more effectively handle the class imbalance than the imputations performed in other ML methods. However, the low sensitivity problem was absent in datasets 2a and 2b, as the number of positives and negatives were equivalent (Table 1).

### Comparison of internal performances of the XGBoost model with published reports

The internal performances of the XGBoost model were compared with reported methods from the published literature^9 11^. The results from the XGBoost model outperformed the published reports (Table 2 and Figure 5).

**Table 2:** Internal evaluation of the XGBoost model on different datasets and comparison with published datasets

| Dataset | Sensitivity | Specificity | Accuracy | AUC score | Published AUC score |
| --- | --- | --- | --- | --- | --- |
| 1 | 0.826 | 0.974 | 0.940 | 0.941 |  |
| 1a | 0.875 | 0.925 | 0.918 | 0.967 |  |
| 1b | 0.750 | 0.887 | 0.869 | 0.922 | 0.87 (ref 8) |
| 1c | 1.000 | 0.906 | 0.918 | 0.939 | 0.87 (ref 8) |
| 2a | 0.830 | 0.843 | 0.835 | 0.906 | 0.84 (ref 10) |
| 2b | 0.845 | 0.733 | 0.787 | 0.842 |  |
| 3a | 0.719 | 0.799 | 0.776 | 0.835 |  |
| 3b | 0.784 | 0.733 | 0.746 | 0.842 |  |

**Figure 5:**
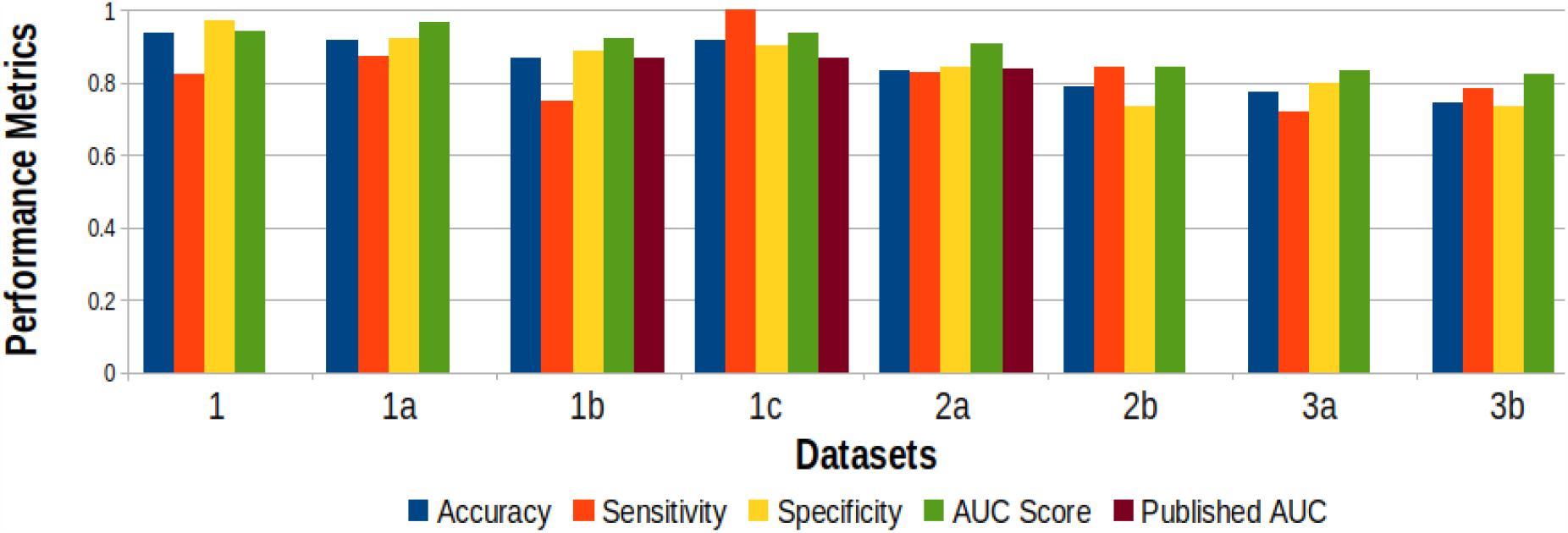
Comparative performances of different datasets trained on XGBoost model. The datasets with published AUC scores are shown in brown bars for the following datasets (1b and 1c)^9^ and 2a^11^.

### Selection of working XGBoost models for external evaluation across the populations

As per the results, the XGBoost model performed the best on dataset 1c, having four hematological parameters. However, the performance of the XGBoost model on dataset 1b, having fourteen hematological parameters, was comparable to that of dataset 1c, with a slightly lower AUC Score (0.94 versus 0.92). Based on these observations, we hypothesize both four-parameter and fourteen-parameter models as the working ML models for COVID-19 testing and blind predictions across different populations. Although the internal performances were the best with datasets 1a and 1c, the overfitting of the data due to small sample sizes was an issue, as discussed above. Hence, we selected two other XGBoost models with four and fourteen parameters obtained from datasets 2b (Italy) and 3b (Brazil), *albeit* with a slightly lowered AUC score of 0.842 in both cases. These two were the final working models (training dataset) for external evaluation.

### External evaluation of XGBoost models with four hematological parameters across Italian and Brazilian populations

External evaluation for the four-parameter model was performed on the test dataset 1c from Brazil. Note that the training dataset 2b was from Italy. The sensitivity was 0.81 with a lower specificity value; the AUC score was 0.69 (Table 3a). For the first time, an ML model was trained on one ethnic group and tested on another ethnic group with reasonably good performance.

**Table 3:** External evaluation of XGBoost algorithm based on a) 4-hematological features and b) 14-hematological features trained and tested across different datasets.

| Training set/test set | Sensitivity | Specificity | Accuracy | AUC Score |
| --- | --- | --- | --- | --- |
| Dataset 2b<br>(Italian) /<br>dataset 1c<br>(Brazilian) | 0.81 | 0.45 | 0.50 | 0.69 |

b)
| Training set/test set | Sensitivity | Specificity | Accuracy | AUC Score |
| --- | --- | --- | --- | --- |
| Dataset 3b<br>(Brazilian) /<br>dataset 1b<br>(Brazilian) | 0.55 | 0.90 | 0.85 | 0.86 |

### External evaluation of XGBoost models with fourteen hematological parameters within the Brazilian populations

The fourteen-parameter XGBoost model was trained on dataset 3b (n=12105) and tested on dataset 1b (n=602), both from Brazilian populations. However, the samples in these two datasets were from different time points; hence those can be considered independent data sources. The AUC score for this prediction was 0.86 (Table 3b). These results were better than the performance for the four-feature XGBoost model across the populations. There could be multiple reasons for the better performance of the fourteen-feature model over the four-feature model, a) the larger size of the training dataset, b) training and prediction data obtained from the same demographic location, that is, Brazil, and c) combination of more number of features with a larger dataset, presumably, yields to a better result.

### Blind prediction of XGBoost models with four hematological parameters on West European populations

To further validate the efficacy of the working models, we have considered one more dataset from published literature with thirty-seven features, including the data points along different stages (time points) of COVID-19^13^. The dataset was from the literature without preprocessing (no feature, records, or data points removed). According to the source authors, two distinct stages of COVID-19 patients, W.E.-*early* and W.E.-*advanced*. Distributions of four hematological parameters across the datasets, 1c, 2b, W.E.-*early*, and W.E.-*advanced*, were compared (Figure 6). The distributions were almost the same across all the datasets for Leukocytes and platelets. For eosinophils and monocytes, the distributions for datasets 2b and W.E.-*early* are very similar. Moreover, distributions across datasets 1c and W.E.- *advanced* were similar for the same features. The external performance of the model on W.E.- *early* dataset (0.65) was high compared to that on W.E.-*advanced* dataset (0.52) (Table 4). To note, W.E.-*early* and W.E.-*advanced* datasets contain information only from COVID-19 patients and no negative controls. Hence, only the sensitivity metric was reported (Table 4).

**Table 4:** Blind prediction of XGBoost model trained on dataset 2b and tested on W.E.-*early* and W.E.-*advanced* datasets. The *early* and *advanced* datasets contain only COVID-19-positive patient results; no negatives were available. Hence, only sensitivity values reported

| Training set/test set | Sensitivity |
| --- | --- |
| Dataset 2b/ *W.E.-early* | 0.65 |
| Dataset 2b/*W.E.-advanced* | 0.52 |

**Figure 6:**
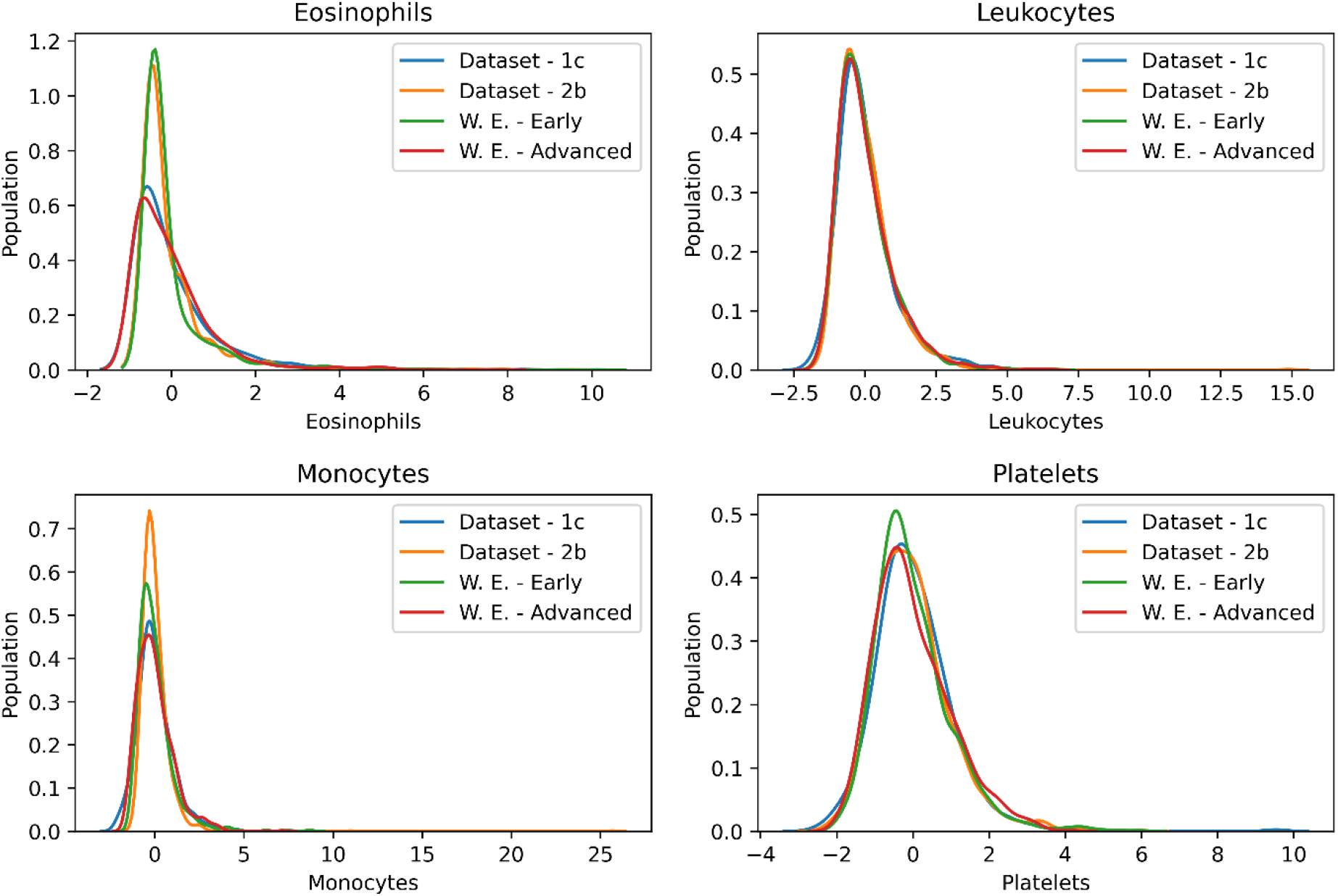
Distributions of four hematological parameters across four different datasets (two training datasets – Dataset 1c and Dataset 2b and two test datasets –*early* and *advance*). The hematological parameters are – a) platelet, b) leukocyte c) eosinophil and d) monocyte. These distributions indicate the proximity of the individual test datasets to the training datasets

### Deployment of Prediction server

We deployed a web server where two sets of inputs are acceptable for binary COVID-19 prediction, i) four hematological parameters (leukocyte, monocyte, eosinophil, and platelet count) and ii) fourteen-parameter models in the following URL link, https://covipred.bits-hyderabad.ac.in/home. Different pages on the webserver are shown (Figure 7). The server outputs the COVID-9 results, either positive or negative, with the COVID-19 probability reported in percentage.

**Figure 7:**
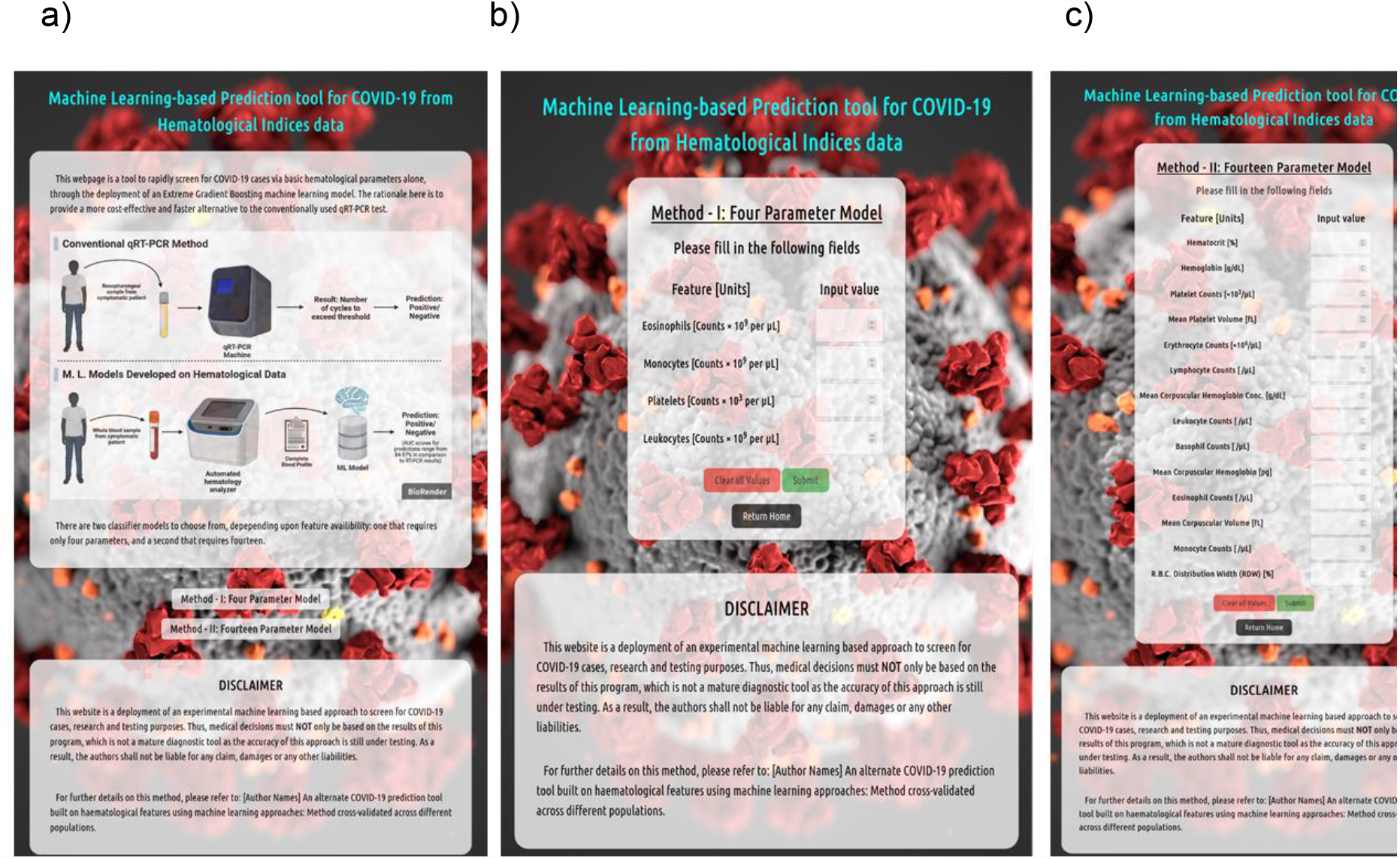
COVID-19 prediction server based on hematological parameters, a) home page b) 4-parameter prediction model and c) 14-parameter prediction model

### Conclusion

Considering the need to develop an alternate protocol for rapid, near-accurate, and cheaper COVID-19 detection techniques, we aimed to externally validate the hematology-based ML prediction reported in the literature with internal evaluation only. We have integrated published clinical records from Brazil, Italy, and West Europe hospitals. The data from Brazil and Italy were classified into eight datasets and trained on seven different ML methods; the XGBoost algorithm was the best. The internal performances of the XGBoost models were better than the published reports on the same datasets. Four and fourteen-parameter XGBoost models were selected for external evaluations. The external performance of the fourteen-parameter XGBoost model trained and tested on the Brazilian dataset was similar to that of the internal performance. However, the external performances of the four-parameter XGBoost model trained on the Italian dataset and tested on a) Brazilian and b) West European datasets were poorer than the previous one. The results promise the utility of these models when trained and tested on the same populations. However, it also warns to use the model, with caution, trained on one population and test on another. The outcome of this work has the potential for an initial screen of COVID-19 based on hematological parameters before qRT-PCR tests. In future work, we aim to train and test those on the Indian population to use at the healthcare centers of India.

## Data Availability

Data sources are mentioned in the manuscript text

https://www.kaggle.com/einsteindata4u/covid19

